# Risk of hypothyroidism in meat-eaters, fish-eaters, and vegetarians: A population-based prospective study

**DOI:** 10.1101/2024.11.25.24317876

**Authors:** Catharina J. Candussi, William Bell, Alysha S. Thompson, Sven Knüppel, Martina Gaggl, Martin Světnička, Jan Gojda, Aedín Cassidy, Cornelia Weikert, Reynalda Córdova, Tilman Kühn

**Affiliations:** Department of Nutritional Sciences, University of Vienna, Vienna, Austria; Center for Public Health, Public Health Nutrition, Medical University of Vienna, Vienna, Austria; Co-Centre for Sustainable Food Systems and The Institute for Global Food Security, Queen’s University Belfast, Northern Ireland, United Kingdom; Unit “Human Study Centre Consumer Health Protection”, Department Food Safety, German Federal Institute for Risk Assessment, Germany; Centre for Research on Diabetes Metabolism, and Nutrition of Third Faculty of Medicine, Charles University, Prague, Czech Republic; Department of Internal Medicine, University Hospital Královské Vinohrady and Third Faculty of Medicine, Charles University, Prague, Czech Republic; Department of Paediatrics, University Hospital Královské Vinohrady and Third Faculty of Medicine, Charles University, Prague, Czech

**Keywords:** Plant-based diets, hypothyroidism, vegan, vegetarian, thyroid, iodine

## Abstract

**Background:** Globally, plant-based diets are gaining in popularity. Whilst their cardiometabolic benefits are well-documented, there is a lack of studies examining the associations between plant-based diets and risk of hypothyroidism.

**Methods:** We analysed data from the UK Biobank cohort. Multivariable Cox proportional hazards regression models were used to estimate hazard ratios (HRs) and 95% confidence intervals (95% CIs) for incident hypothyroidism across vegans, vegetarians, pescatarians, poultry eaters, low meat eaters, and high meat eaters aged 40-69 years. Ancillary to this, we carried out logistic regression analysess to evaluate associations between the diet groups and prevalent hypothyroidism (according to International Classification of Diseases (ICD) codes at baseline.

**Results:** We included 494,437 individuals from the UK Biobank, of which 231,906 followed a high-meat, 236,510 a low-meat, 5,662 a poultry-based, 11,286 a pescatarian, 8,654 a vegetarian, and 419 a vegan diet. During a median (SD) follow-up of 12.7 (± 3.2) years, 24,240 participants developed hypothyroidism. In multivariable Cox regression models without adjustment for body mass index (BMI), none of the diets were significantly associated with the risk of hypothyroidism. However, there was a tendency for a higher risk of hypothyroidism among vegetarians compared to people following a high-meat diet (HR: 1.08, 95% CI: 0.98 - 1.19). After controlling for BMI the association for vegetarians (HR = 1.18, 95% CI 1.07 – 1.30) became stronger and statistically significant. Furthermore, we observed a positive association between a vegetarian (OR: 1.37, 95% CI: 1.14 - 1.63) and a poultry diet (OR: 1.35, 95% CI: 1.10-1.63) with hypothyroidism prevalence.

**Conclusion:** In the present study, we found a moderately higher risk of hypothyroidism among vegetarians, after controlling for BMI, a potential collider. This slightly higher risk of hypothyroidism among vegetarians, requires further investigation, taking iodine status and thyroid hormone levels into account.

## 1. Introduction

Globally plant-based diets are gaining popularity in Western countries. Reasons for this include their potential health benefits, but also animal rights and environmental considerations (1). According to the European ‘Smart Protein’ project approximately 27% of Europeans identify as flexitarian (who try to reduce their animal foods consumption), 4% as pescatarian (who eat fish or shellfish but not any other kind of meat), and 5% as vegetarians (who do not eat meat or fish), while 3% stated that they followed a vegan (who do not eat any kind of animal products) diet (2). Previous research has shown that plant-based diets are associated with lower risks of chronic disease such as type 2 diabetes (3), total cancer (4), cardiovascular disease (5) as well as all-cause mortality (6, 7). Potential health benefits from plant-based diets are particularly noticeable when they are composed of high-quality foods and low in snacks, sugary drinks and ultra-processed foods (6, 8). However, those who consume strict vegan diets may be at risk of critically low intakes of essential nutrients such as n-3 fatty acids, zinc, iron, calcium, or iodine (9-11).

Iodine is a key element for thyroid hormone production. Therefore, an insufficient iodine intake may have detrimental effects on thyroid function, which can lead to goitre, thyroid nodules, and hypothyroidism (12). Additionally, iodine is essential for central nervous system development. Insufficient intake during pregnancy can result in creationism in children which is characterized by impaired growth and intellectual disabilities (13). A sufficient iodine status should be achieved with a daily intake of 150 μg, although a higher intake of 200 μg/day is recommended for pregnant and lactating women (14).

Globally, the leading cause of thyroid disorders, including hypothyroidism, is iodine deficiency (15). In regions where iodine is sufficient, the most common cause of hypothyroidism is Hashimoto’s disease, also known as chronic autoimmune thyroiditis (15, 16). In Europe, the prevalence of hypothyroidism is around 3%, whereby around 5% of hypothyroidism cases may remain undiagnosed (17). Universal salt iodization programs have effectively enhanced iodine intake across numerous countries. In countries such as the United Kingdom (UK), where salt is not routinely iodised, animal products, particularly milk and dairy, as well as seafood, fish and seaweed, contribute significantly to dietary iodine (18-20). However, iodine concentration in milk varies substantially depending on season, soil, farming practices and processing (21). Thus, in the context of a global shift towards predominantly plant-based diets, there is a risk of insufficient iodine intake (22).

Due to the limited research on plant-based diets and hypothyroidism, along with the higher risk of iodine insufficiency for individuals following these diets, the aim of this study was to assess the risk of hypothyroidism among individuals with different types of diet (high-meat eaters, low-meat eaters, poultry eaters, fish-eaters, vegetarians and vegans) using data from the population-based UK Biobank. Our hypothesis was that more plant-based diets would be associated with higher risk due to lower iodine contents, particularly in the UK, where iodine fortification is not mandatory.

## 2. Methods

### 2.1. Study Population

The present analyses are based on data form the UK Biobank, a prospective cohort study comprised of over 500,000 participants aged between 40 and 69 years in the UK. The cohort was conducted between 2006 to 2010 in 22 different assessment centres across England, Scotland and Wales. During the baseline assessment, participants completed a comprehensive touchscreen questionnaire covering details on lifestyle, socioeconomic status and health status as well as 29 items on the frequency of the consumption of certain foods. Additionally, trained professionals conducted physical measurements, and participants provided biological samples. A detailed description of the UK Biobank, including details of the baseline assessment can be found elsewhere (23, 24). Ethical approval for the UK Biobank cohort was obtained from the National Health Service North West Multi-Centre Research Ethics Committee. Written informed consent was received from all participants.

Participants with insufficient data on food frequencies at baseline were excluded from the present analyses (n = 7,748), resulting in a total of 494,437 participants eligible for inclusion in this study. For Cox models, all prevalent hypothyroidism cases (n = 6,633) at baseline were excluded. Furthermore, we had to exclude 284,822 individuals from analysis on iodine intake due to incomplete dietary data (see supplementary Figure 1).

**Figure 1:**
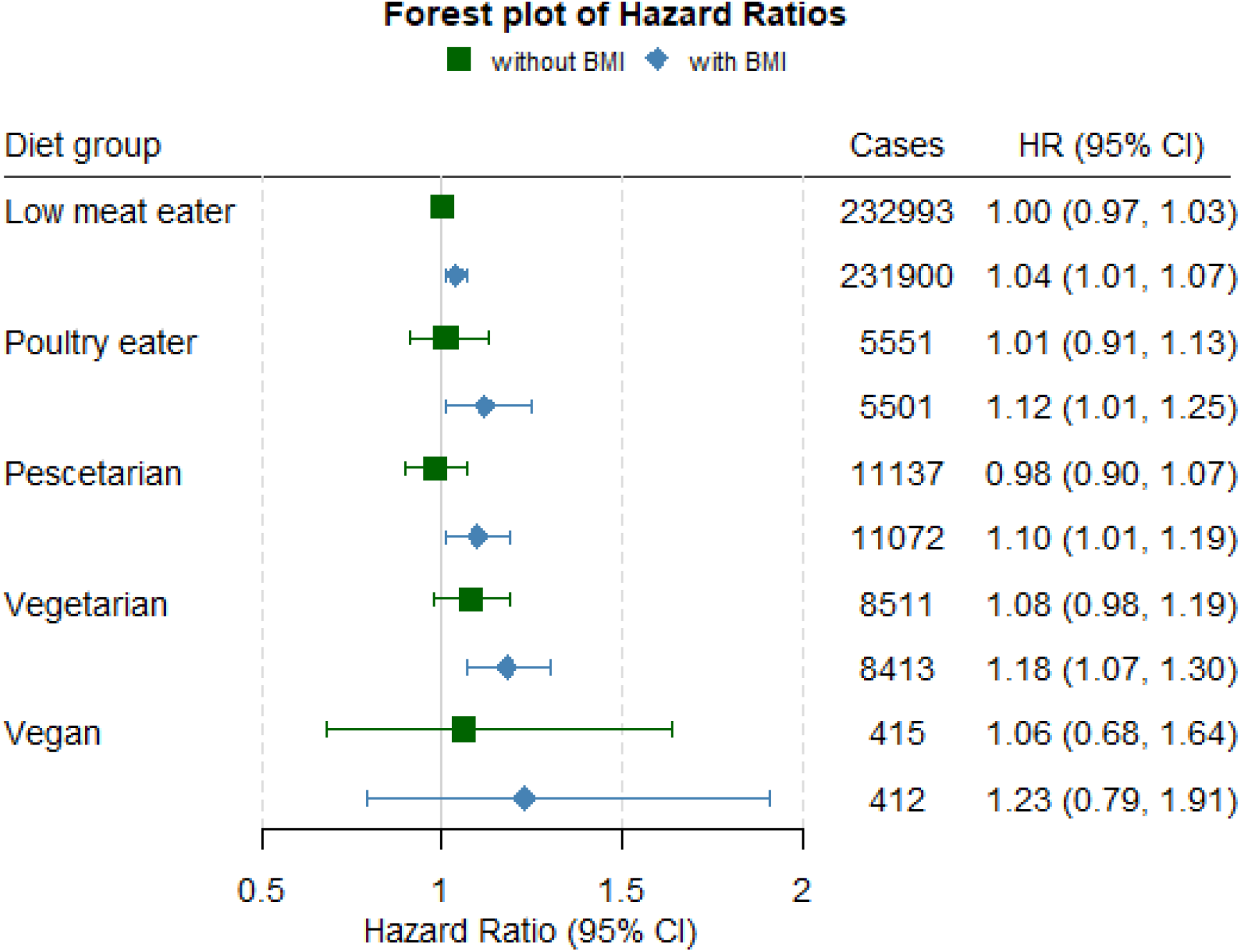
Hazard ratios (HR) and 95% Confidence intervals (95% CI) between diet groups and the risk of hypothyroidism with and without adjustment for body mass index (BMI). All models adjusted for sex, age, income, education, ethnicity, thyroid impairing medication and smoking status.

### 2.2. Dietary Assessment and Categorisation

Study participants were categorised into 6 different diet groups (high meat eater, low meat eater, poultry eater, pescatarian, vegetarian and vegan) according to self-reported food intake in a touchscreen food frequency questionnaire at recruitment, as described by Bradbury et al. and Parra-Soto et al. (25, 26). Thereby the consumption of 29 foods such as processed meat, lamb, pork, poultry, chicken, beef, turkey, oily and non-oily fish as well as cheese and eggs were determined, with frequencies ranging from 0 (never) to 5 (once or more daily). High meat eaters had to consume processed or unprocessed meat (poultry, red meat) more than “5-6 times a week”. Low meat eaters had to eat processed or unprocessed meat at least “less than once a week” but less than “5-6 times a week”. Poultry eaters did not consume red or processed meat, pescatarians did not consume red or processed meat and poultry, vegetarians did not consume red or processed meat, poultry, or fish. Lastly, the vegan group was defined as individuals “never” eating any animal products (see supplementary Figure 2) (25, 26). Detailed data on the consumption of iodine rich foods and iodine intake was obtained by the validated online Oxford WebQ 24-hour dietary questionnaire (27) for a subgroup of 209,615 study participants.

### 2.3. Case Ascertainment

Only potential diet- or iodine related cases of hypothyroidism were used as endpoints for the present study using the UK Biobank’s hospital inpatient data linkage. These cases were identified using the International Classification of Diseases (ICD) codes: the 9th edition (ICD-9) code 244, representing ‘Acquired Hypothyroidism’, and the 10th edition (ICD-10) codes E01.1, E01.8, E02, and E03.9, corresponding to ‘Iodine-Deficiency Related Multinodular (Endemic) Goiter’, ‘Other Iodine-Deficiency Related Thyroid Disorders and Allied Conditions’, ‘Subclinical Iodine-Deficiency Hypothyroidism’, and ‘Unspecified Hypothyroidism’, respectively. Hypothyroidism cases were defined as incident when occurring after baseline assessment and as prevalent when occurring before or at baseline assessment. Nevertheless, a sensitivity analysis was conducted including all cases of hypothyroidism, as identified by the broader ICD-9 codes 240–279, which encompass ‘Endocrine, nutritional, and metabolic diseases’ and ‘Immunity disorders,’ as well as ICD-10 codes E00–E07, which cover ‘Disorders of the thyroid gland.’

### 2.4. Covariate Assessment

Covariate information from baseline assessments was used as follows: Age (continuous: years), sex (categorical: male, female), ethnicity (categorical: Asian, Black, Mixed, White, Unknown), Body Mass Index (continuous: kg/m^2^), average income (categorical: <£18,000, £18000-£30,999, £31,000-£51,999, £52,000-£100,000, >£100,000, unknown), education (categorical: Low, medium, high, unknown), smoking status (categorical: never, previous, current), thyroid impairing medication (categorical: no, yes; including data on the following drugs: Amiodarone, Carbamazepine product, Diethylcarbamazine,, Carbamazepine, Clozapine, Quetiapine, Lithium, Interferon alfa 2a, Interferon alfa 2b, Interferon alfa, Peginterferon alfa 2a, Peginterferon alfa 2b). A table of all covariates and their respective coding and item number from the UK Biobank’s online showcase can be found in Supplementary Table 1.

### 2.5. Statistical Analysis

Descriptive characteristics were assessed across six different diet groups (high meat eaters, low meat eaters, poultry eaters, pescatarians, vegetarians and vegans) and shown as percentages for categorical variables and means ± standard deviations (SD) for continuous variables. To analyse the risk of hypothyroidism across the different diet groups, Cox proportional hazards regression models were used with age as the underlying time variable. The R packages ‘survival’ and ‘survminer’ were used to perform these analyses (28, 29).

Covariates for statistical adjustment were selected following a comprehensive literature search on potential risk factors for hypothyroidism. Based on literature by Chaker et al. (12), Lawton et al. (30), Tonstad et al. (31) and Rizzo et al. (32) we adjusted our Cox models for age, sex, ethnicity, average income, educational level, alcohol intake, physical activity, thyroid impairing medication and smoking status. Additionally, adjusted for alcohol intake and physical activity. However, these potential confounders did not significantly improve model fit and only marginally affected the main associations, which is why we excluded them from the final model. According to the literature, BMI was considered a potential confounder (33, 34). However, since hypothyroidism (possibly also before diagnosis) affects BMI through the physiological influence of thyroid hormones (35, 36), it is important to consider that BMI could be a collider, for which no adjustment should be carried out (see supplementary Figure 3). Therefore, our analyses are presented both with and without adjustment for BMI, the latter to account for potential collider bias. Furthermore, additional sensitivity analyses were conducted accounting for hypothyroidism subtypes restricting models to iodine related hypothyroidism vs. models based on all cases of hypothyroidism (see Case Ascertainment).

In addition to our main prospective analyses, we conducted logistic regression analyses to investigate whether diet groups were associated with prevalent hypothyroidism, adjusting for the same set of confounders. Results were presented as Hazard Ratios and 95% confidence intervals (95% CI) for Cox models, and Odds Ratio (OR) and 95% CIs for logistic regression analyses. The proportional hazards assumption was tested via the Schoenfeld residual test. No violations of proportionality were observed. Results were considered as statistically significant at a two-sided p-values of < 0.05. All statistical analyses were carried out using R (R version 4.3.1) (37).

## 3. Results

### 3.1. Characteristics of the Study Population

Baseline characteristics of the cohort are presented in Table 1. Our analysis included a total of 487,524 participants, 52.8% of whom were female, with a mean age of 56.4 ± 8.1 years. Overall, 47.2% followed a high meat diet, 47.5% a low meat diet, 1.1% a poultry diet, 2.3% a pescatarian diet, 1.7% a vegetarian diet, and >0.1% a vegan diet. During a mean follow-up period of 12.7 ± 3.2 years, 24,233 (5%) participants developed potential iodine related hypothyroidism. The proportion of hypothyroidism cases across the diet groups were as follows: 4.4% among high meat eaters, 5.4% among low meat eaters, 6.1% among poultry eaters, 5.1% among pescatarians, 5.5% among vegetarians, and 4.8% among vegans (see Supplementary Table 2).

**Table 1:** Demographic characteristics of incident hypothyroid cases.

|  | Characteristics | Non-cases (N = 470,197) | Cases (N = 24,240) |
| --- | --- | --- | --- |
| Diet | High meat eaters | 218,932 / 463391 (47%) | 10,144 / 24133 (42%) |
|  | Low meat eaters | 220,253 / 463391 (48%) | 12,599 / 24133 (52%) |
|  | Poultry eaters | 5,208 / 463391 (1.1%) | 338 / 24133 (1.4%) |
|  | Pescatarians | 10,566 / 463391 (2.3%) | 566 / 24133 (2.3%) |
|  | Vegetarians | 8,037 / 463391 (1.7%) | 466 / 24133 (1.9%) |
|  | Vegans | 395 / 463391 (<0.1%) | 20 / 24133 (<0.1%) |
| Sex | Female | 244,757 / 463391 (53%) | 18,855 / 24133 (78%) |
|  | Male | 218,634 / 463391 (47%) | 5,278 / 24133 (22%) |
| Age |  | 56.37 (8.11) | 58.84 (7.39) |
| Ethnicity | White | 437,894 / 463390 (94%) | 22,930 / 24133 (95%) |
|  | Mixed | 2,742 / 463390 (0.6%) | 122 / 24133 (0.5%) |
|  | Asian | 9,949 / 463390 (2.1%) | 616 / 24133 (2.6%) |
|  | Black | 7,217 / 463390 (1.6%) | 211 / 24133 (0.9%) |
|  | Unknown | 5,589 / 463390 (1.2%) | 254 / 24133 (1.1%) |
| BMI (kg/m<sup>2</sup>) |  | 27.34 (4.73) | 28.49 (5.43) |
|  | Unknown | 2,380 | 149 |
| Average Income | Greater than 100,000 | 22,196 / 463391 (4.8%) | 583 / 24133 (2.4%) |
|  | 52,000 to 100,000 | 82,578 / 463391 (18%) | 2,762 / 24133 (11%) |
|  | 31,000 to 51,999 | 104,536 / 463391 (23%) | 4,438 / 24133 (18%) |
|  | 18,000 to 30,999 | 99,903 / 463391 (22%) | 5,562 / 24133 (23%) |
|  | Less than 18,000 | 86,972 / 463391 (19%) | 6,195 / 24133 (26%) |
|  | Unknown | 67,206 / 463391 (15%) | 4,593 / 24133 (19%) |
| Education | High | 219,482 / 463391 (47%) | 9,773 / 24133 (40%) |
|  | Medium | 84,102 / 463391 (18%) | 4,026 / 24133 (17%) |
|  | Low | 151,897 / 463391 (33%) | 9,804 / 24133 (41%) |
|  | Unknown | 7,910 / 463391 (1.7%) | 530 / 24133 (2.2%) |
| Smoking status | Current | 48,841 / 463391 (11%) | 2,417 / 24133 (10%) |
|  | Previous | 159,317 / 463391 (34%) | 9,065 / 24133 (38%) |
|  | Never | 253,658 / 463391 (55%) | 12,553 / 24133 (52%) |
|  | Unknown | 1,575 / 463391 (0.3%) | 98 / 24133 (0.4%) |
| Physical activity | High | 111,479 / 463391 (24%) | 5,303 / 24133 (22%) |
|  | Moderate | 183,100 / 463391 (40%) | 8,601 / 24133 (36%) |
|  | Low | 65,740 / 463391 (14%) | 3,564 / 24133 (15%) |
|  | Unknown | 103,072 / 463391 (22%) | 6,665 / 24133 (28%) |
| Thyroid impairing drugs |  | 1,611 / 463391 (0.3%) | 253 / 24133 (1.0%) |

Abbreviation: BMI, body mass index (calculated as weight in kilograms divided by height in meters squared). Categorical variables are expressed as the number of cases divided by the total number of observations (n/N), followed by the percentage in parentheses: n/N (%). Continuous variables are expressed as the arithmetic mean and standard deviation (mean ± (SD)).

The consumption of iodine rich foods across diet groups among the 209,615 participants with 24-hour dietary data is shown in Supplementary Table 3. Intakes of dairy and cheese were similar among high and low meat eaters as well as poultry eaters, ranging from 0.5 to 0.65 servings per day. Pescatarians and vegetarians reported a higher dairy and cheese intake (0.71-0.94 servings/day), while vegans reported the lowest intakes (0.21 servings for dairy and 0.17 for cheese). Fish consumption was highest among pescatarians and poultry eaters (0.47 servings/day), while vegetarians and vegans reported a very low consumption (0.01 servings/day). Poultry intake was highest in high and low meat eaters (0.32-0.34 servings/day), while pescatarians and vegetarians reported to consume very little to none. Red meat consumption was most prevalent among high meat eaters (1.12 servings/day) and moderate in low meat eaters (0.72 servings/day). Poultry eaters, pescatarians, vegetarians, and vegans consumed minimal amounts of red meat (0.01 to 0.14 servings/day). Those following a high meat, low meat, poultry, or pescatarian diet had average iodine intakes of around 200 μg/day (range: 217-204 μg/day). In contrast, those following a vegetarian or vegan diet had mean iodine intakes of 164 (±68) μg/day and 93 (±45) μg/day, respectively (see Supplementary Table 4). As seen in Supplementary Table 5 about 92% of vegans and 44% of vegetarians did not meet a sufficient daily iodine intake of 150 μg.

Overall, individuals who developed hypothyroidism were more likely to be female (78%), had a higher BMI (28.5), and had a lower income (see Table 1). Similar patterns were observed for prevalent hypothyroidism cases (n = 6,620), with the majority being females (84%) and from lower income groups at baseline (see Supplementary Table 6).

### 3.2. Diets and Risk of Hypothyroidism

As shown in Figure 1, without BMI adjustment none of the diet groups were significantly associated with hypothyroidism risk, although a marginal association was observed for vegetarians (HR = 1.08 95% CI 0.98 – 1.19). After controlling for BMI the associations for low meat eaters (HR = 1.04 95% CI 1.01 – 1.07) poultry eaters (HR = 1.13 95% CI 1.01 – 1.25), pescatarians (HR = 1.10 95% CI 1.01 – 1.19), and vegetarians (HR = 1.18 95% CI 1.07 – 1.30) became stronger and statistically significant. In sensitivity analyses including non-iodine related hypothyroidism cases, we observed similar results across all diet groups (see supplementary Figure 4).

Logistic regression analyses on vegetarian diet and prevalent hypothyroidism showed a multivariable-adjusted OR (95% CI) of 1.20 (1.01 - 1.43) and 1.37 (1.14 – 1.63), respectively, after controlling for BMI (Table 2). Furthermore, greater odds for poultry eaters (OR = 1.35 95% CI 1.10 – 1.63) we observed after BMI adjustment.

**Table 2:**
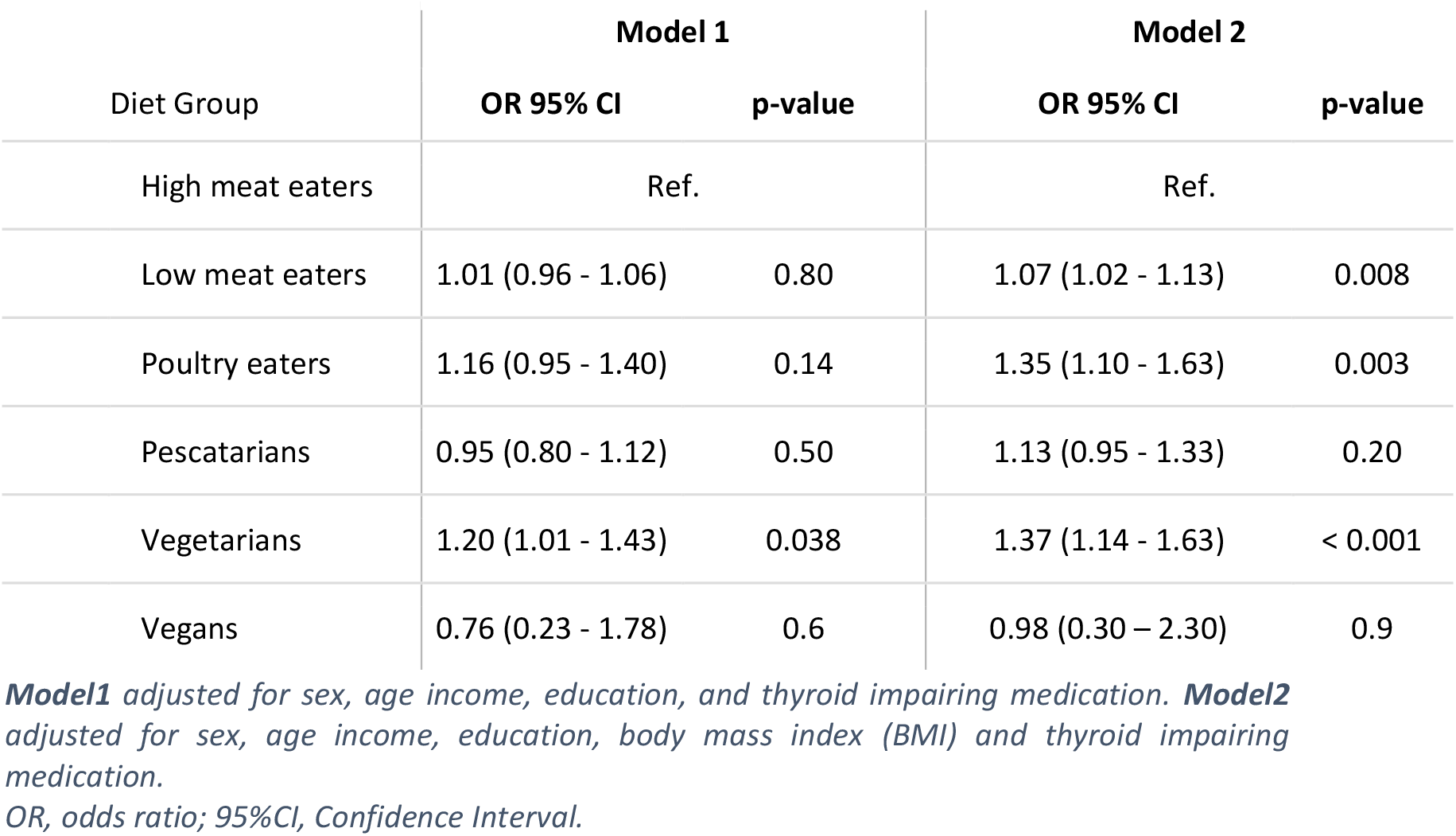
Odds ratios of prevalent hypothyroidism (n = 6,633) across diet groups in 494,144 UK-Biobank participants.

### 4. Discussion

In the present study, we observed no statistically significant associations between diets lower in animal products and hypothyroidism risk in multivariable models not adjusted for BMI. After adjusting for BMI, the associations for low meat eaters (HR = 1.04 95% CI 1.01 – 1.07), poultry eaters (HR = 1.12 95% CI 1.01 – 1.25), pescatarians (HR = 1.10 95% CI 1.01 – 1.19), and vegetarians (HR = 1.18 95% CI 1.07 – 1.30) became slightly stronger in magnitude and statistically significant. Similarly, analyses on diet types and hypothyroidism cases at recruitment (prevalent cases) showed greater odds among vegetarians (OR= 1.37 95% CI 1.14-1.63) and poultry eaters (OR = 1.35 95% CI 1.10 – 1.63) compared to high meat eaters, while no associations between other diet types and hypothyroidism were observed.

Given the influence of BMI on our results, it is crucial to consider whether BMI constitutes a collider or a confounder regarding the association between type of diet and hypothyroidism. A confounder affects both the exposure and the outcome (38). A recent Mendelian randomization analysis by Qiu et al. indicates that BMI may affect hypothyroidism, the outcome in our study, as genetically predicted BMI was associated with higher risk of hypothyroidism (39). In turn, genetically predicted hypothyroidism was not associated with higher BMI. Considering the associations between BMI with both diet type and hypothyroidism in our study, adjustment for BMI as a confounder would be necessary. However, it seems likely that the type of diet in our study is a determinant of BMI rather than vice versa, given lower calorie intake among people following plant-based diets (40). In addition, a secound Mendelian randomization by Zhou et al. did not show an association between genetically predicted BMI and hypothyroidism (41). Alternatively, BMI may introduce collider bias, as it is influenced by both diet, through lower caloric intake among people following a more plant-based diet (40), and hypothyroidism, through the metabolic changes associated with the condition, even in its subclinical from with potential effects prior to diagnosis (42). While collider bias seems plausible, we can only speculate that undiagnosed hypothyroidism or impaired thyroid function affected BMI in our study, as no data on thyroid health prior to BMI measurements were available. Thus, despite studies on the occurrence of unrecognised subclinical hypothyroidism among people with higher BMI, the temporality of increases in BMI and the occurrence of hypothyroidism in our study remains unclear (17, 43).

In a similar study from the United States, Tonstad et al. analysed data of 97,000 American adults enrolled in the Adventist Health Study II (AHS-II). Study participants following a vegan diet did not show a higher risk of hypothyroidism compared to omnivores (OR 0.89, 95% CI: 0.78-1.01) (8). By contrast, a higher risk was observed for vegetarians (OR 1.09, 95% CI: 1.01-1.18) for hypothyroidism, which is in line with the present findings. Of note, the analyses of Tonstad et al. were adjusted for BMI. To our knowledge there are no other studies on diet types in relation to hypothyroidism, although two studies, in which dietary patterns were analysed, are somewhat consistent with our findings. Alkhatib et al. observed in a cohort from three cycles of National Health and Nutrition Examination Surveys (NHANES) that people following a ‘fat–processed grains–sugars–meats’ or a ‘oils–nuts–potatoes–low-fat meats’ dietary pattern had lower risk of hypothyroidism compared to those with ‘fruits–whole grains–vegetables–dairy’ pattern (44). In another cohort study from South Korea participants were grouped by four dietary patterns (Korean balanced diet, plant-based diet (PBD), Western-style diet (WSD) and rice-based diet). The study found that individuals with a high polygenic risk and a high plant-based diet score had a risk of hypothyroidism (45).

A systematic review and metanalysis including 11 studies (n = 4421) showed that a vegan diet is associated with lower intakes of iodine and lower median urinary iodine concentration (12.2– 44.0 μg/l) compared to omnivores (46). According to the world health organisation, optimal urinary iodine levels in adults range between 100–200 μg/l (47). A sufficient iodine status should be achieved with a daily intake of 150 μg, although a higher intake of 200 μg/day is recommended for pregnant and lactating women (14). Especially in regions where universal salt iodisation programmes are absent or voluntary such as the UK, studies suggest that vegetarian and vegan diets are not appropriate to meet dietary recommendations and may increase the risk for iodine deficiency (48, 49). This is also reflected by our results, with the majority of individuals following an omnivorous diet demonstrating adequate iodine intake (>150 μg), compared to 55.6% of vegetarians and just 7.8% of vegans.

The present results on iodine intake may seem surprising, as dairy products that are consumed by vegetarians are the main source of dietary iodine in the UK (50). However, there is wide variability in iodine levels in milk which depend on a range of factors including the farming practice used (21). Overall cow’s milk naturally contains a low concentration of iodine but becomes a good source of this nutrient due to typical farming practices such as using ionized disinfectant or adding iodine salts to cattle feed (51). According to a market survey data conducted in the UK, around 20% of plant-based dairy products are fortified with iodine and iodine levels in cow milk were found to be ten times higher than those in plant-based alternatives (52). Iodine intakes were lower among vegetarians in our study compared to meat eaters and pescatarians, and the differences in average iodine intakes are potentially attributable to omitting fish and meat. Vegans, who had substantially lower iodine intakes compared to all other groups in our study, did not have a higher risk of hypothyroidism, although analyses among vegans were in our study were restricted to a very small number of vegans. Our finding of an only slightly greater risk of hypothyroidism among vegetarians could be partly due to the thyroid gland’s ability to adapt to mild iodine deficiency, sustaining normal thyroid hormone synthesis (53). Henjum et al. investigated that thyroid hormone concentrations among vegans and vegetarian remain normal, despite the fact that both diets are associated with lower urinary iodine concentrations (54). Extensive epidemiological research indicates that prolonged thyroid stimulation, resulting from this adaptive response, promotes thyroid enlargement. During this phase of follicular cell proliferation, there is an increased likelihood of mutations, which can lead to multifocal autonomous growth and subsequent thyroid dysfunction (53, 55). Thus, further research on plant-based diets and thyroid health from population-based studies in needed to monitor potential adverse effects.

One limitation of this study is its observational design, as residual or unmeasured confounding cannot be ruled out. Despite a substantial number of hypothyroidism cases during follow-up, our analyses were not well-powered to detect moderate associations among vegans due to the relatively low number of individuals in this group. Additionally, primary care data was not available to identify hypothyroidism cases in this study. Misclassification of diet groups is also a limitation, as participants may have altered their diet during the follow-up or underreported their intake. This is reflected by the lower amounts of meat and fish consumption reported during 24-hour dietary assessments by some individuals classified as vegans or vegetarians according to the baseline food frequency questionnaire. Such misclassification may have led to a potential underestimation of the risk estimates in our study. Data on iodine intake were only available for a subsample of the population and there were no data on iodine status or iodine hormone levels. No data on iodine supplementation or fortified vegan products was available leading to a potential underestimation of iodine intake, although iodine intake data is consistent with data on iodine status among people following plant-based diets from previous studies (46, 54). Moreover, the dietary data collected from the baseline touchscreen questionnaire did not include total energy intake, so energy could not be included as a potential confounder in the multivariable models. Finally, the UK Biobank cohort represents a population with a healthier risk profile compared to the general population and includes predominantly British participants, most of whom are of white European ancestry (56, 57). This may limit the generalizability of the findings to other populations.

Overall, the findings from our study indicate that a vegetarian diet may be associated with a moderately higher risk of hypothyroidism. This finding is consistent with findings from a previous study, and warrants further investigation in studies with data on iodine status and thyroid function prior to diagnosis.

## Data Availability

All data produced in the present study are available upon reasonable request to the authors

